# Examining risk factors for weight change during midlife: A Mendelian randomization study

**DOI:** 10.1101/2021.07.21.21260895

**Authors:** Grace M. Power, Jessica Tyrrell, Apostolos Gkatzionis, Si Fang, Jon Heron, George Davey Smith, Tom G. Richardson

## Abstract

**Background:** Maintaining a healthy weight across adulthood reduces morbidity and mortality risk in later life. This study aimed to examine factors contributing to weight change over a one-year interval in midlife. While conventional epidemiological analyses have reported risk factors associated with weight change, biases such as confounding present challenges when inferring causality.

**Methods:** Conventional observational analyses were used in addition to a one-sample Mendelian randomization (MR) approach to estimate the genetically predicted effects of four exposures (alcohol consumption, smoking intensity, educational attainment and Alzheimer’s disease liability) on weight change (mean age: 56.9 years) using data from 329 531 participants in the UK Biobank.

**Results:** One-sample MR indicated strong evidence that Alzheimer’s disease liability increased the odds of weight loss whilst conventional analyses reported little evidence of this. In MR and conventional epidemiological analyses, higher educational attainment was associated with maintaining a steady weight. In addition, higher alcohol consumption was associated with weight gain in conventional analyses only. Finally, whilst conventional analyses showed that smoking heaviness was associated with weight gain, the converse was supported by MR, which indicated strong evidence that smoking heaviness reduced the odds of weight gain.

**Conclusions:** Our findings highlight important risk factors for weight change in midlife and emphasise the public health importance of evaluating the dynamic changes to body weight within a causal inference setting.

## Background

Age-related changes in weight and body composition can have major health implications (1–5). Whilst weight gain in adulthood has been associated with increased risk of cardiometabolic disease, including type 2 diabetes and coronary heart disease (1, 5), weight loss in midlife may be an early indicator of dementia and increased mortality risk in later life (2–4). Mendelian randomization (MR) is a technique that can be used to generate evidence about the causal effects of modifiable exposures on outcomes by exploiting the quasi-random assortment of genetic variants associated with an exposure of interest (6, 7). As an individual’s genotype is established at zygote formation, genetic variation is robust to reverse causation and confounding is substantially less evident than in conventional epidemiological studies (6, 7). Often MR studies investigating weight, however, employ a single cross-sectional measure of body mass index (BMI) overlooking the dynamic feature of body weight change over time (3). To aid in the prevention of long-term consequences resulting from weight change in midlife, gaining a comprehensive understanding of the genetically predicted effects of potential risk factors on weight change during this lifecourse interval, is of key public health importance.

Lifestyle risk factors, including increased alcohol consumption and smoking intensity, have been shown to associate with weight change (8–10). Alcohol has previously been reported to increase weight as a consequence of excess calorie intake (10). However, evidence is varying, with several studies also indicating absent or weak associations between alcohol consumption and weight gain (11, 12). Findings concerning the relationship between smoking and weight change are additionally conflicting (8, 9). Such inconsistencies are likely due to the influence of other lifestyle factors since physical inactivity, unhealthy diet and alcohol intake are positively correlated with both adiposity and smoking behaviour (13).

Another marker often shown to be associated with weight change is education. Higher educational attainment was shown to be associated with a reduced increase in weight gain during adulthood in a large multicentre prospective cohort study conducted between 1992 to 2000 across Europe (n=361,467) (14). Several studies have also reported an inverse association between education and changes in body mass index (BMI) measured across the lifecourse (14, 15). Furthermore, a 2013 systematic review comprising 289 associational studies assessing educational attainment and obesity identified an inverse relationship in studies from higher-income countries (16). Evident within this review, however, was a sparsity of investigations that adequately adjusted for all potential confounders, highlighting the inherent issues of residual confounding within conventional observational research (6, 7).

Whilst it is well documented that weight loss occurs more frequently in patients with Alzheimer’s disease than amongst their cognitively normal counterparts, there is some evidence that weight loss manifests before Alzheimer’s disease (17). MR has been extended to evaluate the genetically predicted effects of disease liability on outcomes (18), which includes diseases that typically have a late-onset in the lifecourse, such as dementia. Since latent liability to dementia can be thought of as probabilistically underlying the binary diagnosis measure, results are interpreted on the liability scale (19). Within the data employed in this study, participants are likely to be at different stages of disease development and some who would never develop disease may still show influences of liability, emphasising the need for interpretation to be the effect of disease liability.

This study aims to estimate the effects of Alzheimer’s disease liability, educational attainment, alcohol consumption and smoking on self-reported weight change over a one-year interval in midlife, using data on participants from the UK Biobank study (20). Initially, conventional epidemiological analyses will be conducted. MR analyses will subsequently be applied. Whilst individuals considered overweight are shown to be more likely to under-report weight (16), the discrepancy between self-reported weight change compared to measured weight change has previously been shown to be minor (21). Even so, this investigation seeks to validate the self-reported weight change measure used in this study, by examining the relationship between self-reported weight loss and weight gain and all-cause mortality in later life, an outcome shown to result from unintentional weight changes (3–5).

## Methods

### Data resources

Data were collected from the UK Biobank study; a population-based cohort aged between 40-69 years at recruitment, enrolled in the United Kingdom between 2006 and 2010 (20). In depth details on data collection within UK Biobank during clinic visits and through electronic health records, as well as genotyping, quality control and imputation have been described in detail previously (22). The UK Biobank study have obtained ethics approval from the Research Ethics Committee (REC; approval number: 11/NW/0382). The primary data from the UK Biobank resource that support the findings of this study are accessible upon application (https://www.ukbiobank.ac.uk/).

All observational exposure data were self-reported and collected during a single wave between 2006 and 2010 (Supplementary Table S1). Smoking intensity was assessed by number of cigarettes per day. Alcohol intake was assessed by units per day, which was calculated from the NHS estimates of pure alcohol consumption (23). Weekly and monthly average intake was converted to daily intake by dividing by 7 and 30, respectively. Educational attainment was derived as a continuous score using the criteria described previously by the Social Science Genetic Association Consortium (SSGAC) consortium (24). This is measured as the number of years of schooling that individuals completed. Family history of Alzheimer’s disease was analysed as a binary variable indicating either or both parents reportedly having Alzheimer’s disease, as well as subsequent analyses of mothers and fathers separately. The outcome variable used in this investigation was obtained from participants being asked the question “Compared with one year ago, has your weight changed?” with the response options “Yes – lost weight”, “No – weight about the same” and “Yes – gained weight”. Whilst a popular approach to carrying out analyses on a categorical outcome with more than two groups is to implement a multinomial regression, this method may not be able to recover causal effects due to collider bias (25). The outcome of interest was therefore separated into two binary dependent variables; weight loss compared to staying the same and weight gain, coded 1 and 0, respectively, and weight gain compared to staying the same and weight loss, coded 1 and 0, respectively. In this instance, no individuals were dropped helping to mitigate against bias. Complete data were available on 329 531 participants of European ancestry in the UK Biobank. Missing observations were excluded from analyses. The variable indicating all-cause mortality, taken from the death register and accessible via the UK Biobank, was used to validate our outcome variable, since self-reported unintentional weight change has previously been shown to strongly associate with mortality (3–5).

### Genetic risk scores

In order for a genetic variant to be considered an instrumental variable for an exposure in MR analyses, the instrumental variables used must i) associate with the exposure of interest conditional on the other exposures (the ‘relevance’ assumption), ii) not affect the outcome except through the exposures (the ‘exclusion restriction’ assumption) and iii) be independent of all confounders, both observed and unobserved, of any of the instrumental variables and outcome (the ‘exchangeability’ assumption) (6, 7). Instruments for each exposure were constructed as mr genetic risk scores coded additively in UK Biobank using the software PLINK (26). Genetic variants robustly associated with alcohol intake (27), educational attainment (28) and Alzheimer’s disease (29) were identified from large-scale genome-wide association studies which did not include the UK Biobank study. Variants were selected based on P<5×10^-8^ and r^2^<0.001 using a reference panel of Europeans from the 1000 genomes project phase 3 to ensure they were independent (30). Genetic influence on smoking is poorly captured by the observed variable indicating number of cigarettes smoked per day since the largest effect variant relates to other aspects of smoking behaviour that influence biological exposure (31). Thus, for smoking intensity, we used a single variant as our instrument (rs16969968) located at the CHRNA5 locus. This genetic variant relates to lung cancer risk in smokers, though not in non-smokers and has been shown to be robustly associated with smoking heaviness amongst smokers (32).

### Statistical analysis

Logistic regression analyses were undertaken to estimate the association of each of the four exposures with weight loss (compared to staying the same and weight gain) and weight gain (compared to staying the same and weight loss). All analyses were adjusted for age, sex and the top ten principal components to account for residual population stratification in the UK Biobank study.

Univariable MR analyses were conducted using individual-level data by applying logistic regression with the genetic instruments described above on the weight change variables. Using one-sample MR, as opposed to two-sample MR, allowed us to remove participants who had reportedly never smoked or who do not drink. This ensured analyses were conducted accurately on appropriate populations: ever smoked (n=150 799) and ever drunk alcohol (n=312 923). MR analyses were adjusted for the same covariates as in the conventional epidemiological analysis. The principal components were derived from the UK Biobank genetic data (20).

Previous evidence has found that the effect of ever smoking on BMI weakens as time since smoking cessation increases (33). Therefore, MR analyses look at the relationship between the genetic instrument used for smoking heaviness and weight change were also completed after stratifying ever smokers into current (n=33 094) and previous (n=117 705) smokers. Furthermore, conducting MR analyses in current smokers ensured comparisons could be made between MR and conventional epidemiological estimates, since the observed smoking intensity variable was measured by number of cigarettes smoked per day and is therefore collected for current smokers only.

#### Collider bias

Inverse Probability Weighting (IPW), an approach that uses weights to attempt to make selected participants a representative sample of the study population, has been shown to ameliorate collider bias in MR studies (34, 35). We used IPW to account for both (i) selection on smoking and alcohol use and (ii) stratification by smoking status. As illustrated in Figure 1, the exclusion of never-smokers (or never-drinkers) opens a pathway between the genetic instrument used and the outcome, weight change, via both measured and unmeasured confounders (Figure 1A). Likewise, stratification within the ever smokers’ group to investigate relationships within previous and current smokers may also induce bias (Figure 1B). The use of IPW was restricted to MR analyses in our paper, since data were not available to conduct the same analyses using conventional methods.

**Figure 1:**
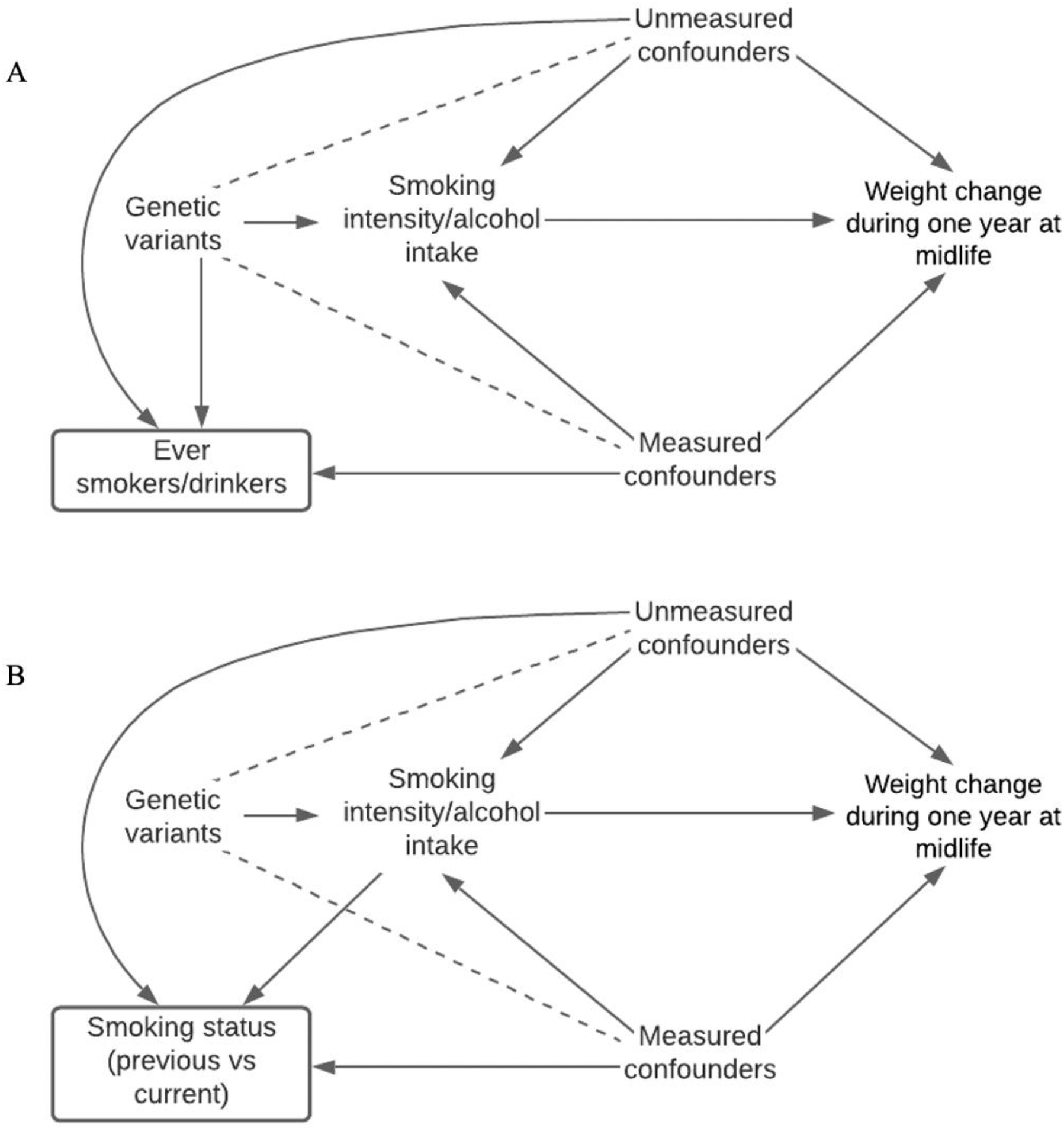
Directed acyclic graphs indicating potential collider bias. Collider bias could be encountered in this study when selecting participants based on smoking and alcohol drinking status in the UK Biobank (A) and collider bias could be encountered in this study when stratifying on participants’ smoking status in the UK Biobank (B).

IPW weights were derived from a logit model estimated on the whole population. The logit model included the collider variable (never vs ever smokers) as the binary outcome, and risk factors for smoking initiation as independent variables. These weights were then included in the analysis assessing the effect of the genetic instrument used for smoking intensity on weight change in ever smokers. A similar model was fitted to assess the association between alcohol consumption and weight change in ever drinkers. A second logit model was estimated within the ever (previous vs current) smoking population. This included risk factors for current smoking. Weights from this second model were multiplied by the previous weights described above. These new weights were then included in the stratified analysis assessing the effect of the genetic instrument for smoking intensity on the weight change variables within previous and current smokers. This technique allows for each selected participant to account for themselves and for those with similar characteristics who were not selected. Reduced sample sizes were a result of including additional variables in our models as data on all variables were necessary to conduct this complete-case analysis.

#### Validation

Self-reported unintentional weight loss over a one-year interval in adulthood has previously been shown to strongly relate to mortality in later life (4). To validate the self-reported weight change variable employed within this study, we conducted logistic regression analyses to calculate the association between weight loss compared to remaining the same weight and weight gain compared to remaining the same over a one-year period in midlife, and all-cause mortality. Since our weight change measures were considered independent in this context, dropping those that reported weight gain and weight loss, respectively, from these variables would not induce collider bias. Analyses were adjusted for age, sex and the top ten principal components.

#### Sensitivity analyses

We conducted sensitivity analyses using MR to apply the pleiotropy robust weighted median and MR-Egger methods using the ‘TwoSampleMR’ R package (36, 37). This was computed for the exposures measuring alcohol consumption, educational attainment and Alzheimer’s disease liability to examine the robustness of results to horizontal pleiotropy; the phenomenon where genetic variants influence multiple traits or disease outcomes via independent biological pathways. Pleiotropy robust methods were not undertaken for smoking intensity due to only using a single SNP instrument at the *CHRNA5* locus. A two-sample MR leave one out analysis was additionally undertaken, when estimating the effects of Alzheimer’s disease, by removing the ‘rs12972156’ genetic variant, located at the *APOE* locus. This was to ensure that the ‘rs12972156’ genetic variant did not distort the overall results. There may be some potential for bias due to sample overlap in these analyses. We conducted further analyses to provide evidence that the weight change variable used in this study was indicative of unintentional weight change. For this, we identified a variable in the UK Biobank showing major dietary changes in the last 5 years (n=334 212). This comprised three groups: ‘No’ (61.6%; 205 858/334 212), ‘Yes, because of illness’ (10.5%; 31 112/334 212) and ‘Yes, because of other reasons’ (27.9%; 93 242/334 212). Within the group that answered ‘No’, we reran our analyses to generate estimates for risk factors of interest on weight change during midlife.

Forest plots in this paper were generated using the R package ‘ggplot2’ (38). These analyses were undertaken using R (version 3.5.1).

## Results

The study sample used comprised of 177 389 (53.8%) females and 152 142 (46.2%) males with a mean (SD) age of 56.9 (8.0) years. Weight loss was reported by 50 262 (15.3%), weight gain by 92 057 (27.9%) and staying the same weight by 187 212 (56.8%) participants over a one-year interval during midlife.

In MR analyses, there was strong evidence that a 1-SD increase in Alzheimer’s disease liability increased odds of weight loss (Odds ratio (OR), 95% CI: 1.023, 1.013 to 1.033, P=2.67×10^-6^) whilst in conventional observational analyses, there was very little evidence of this effect (OR, 95% CI: 1.019, 0.991 to 1.049, P=0.188). Tables 1 and 2 and Figure 2A show educational attainment and alcohol consumption reduced odds of weight loss, in both conventional observational and MR estimates, and smoking intensity conferred increased odds of weight loss.

**Table 1:**
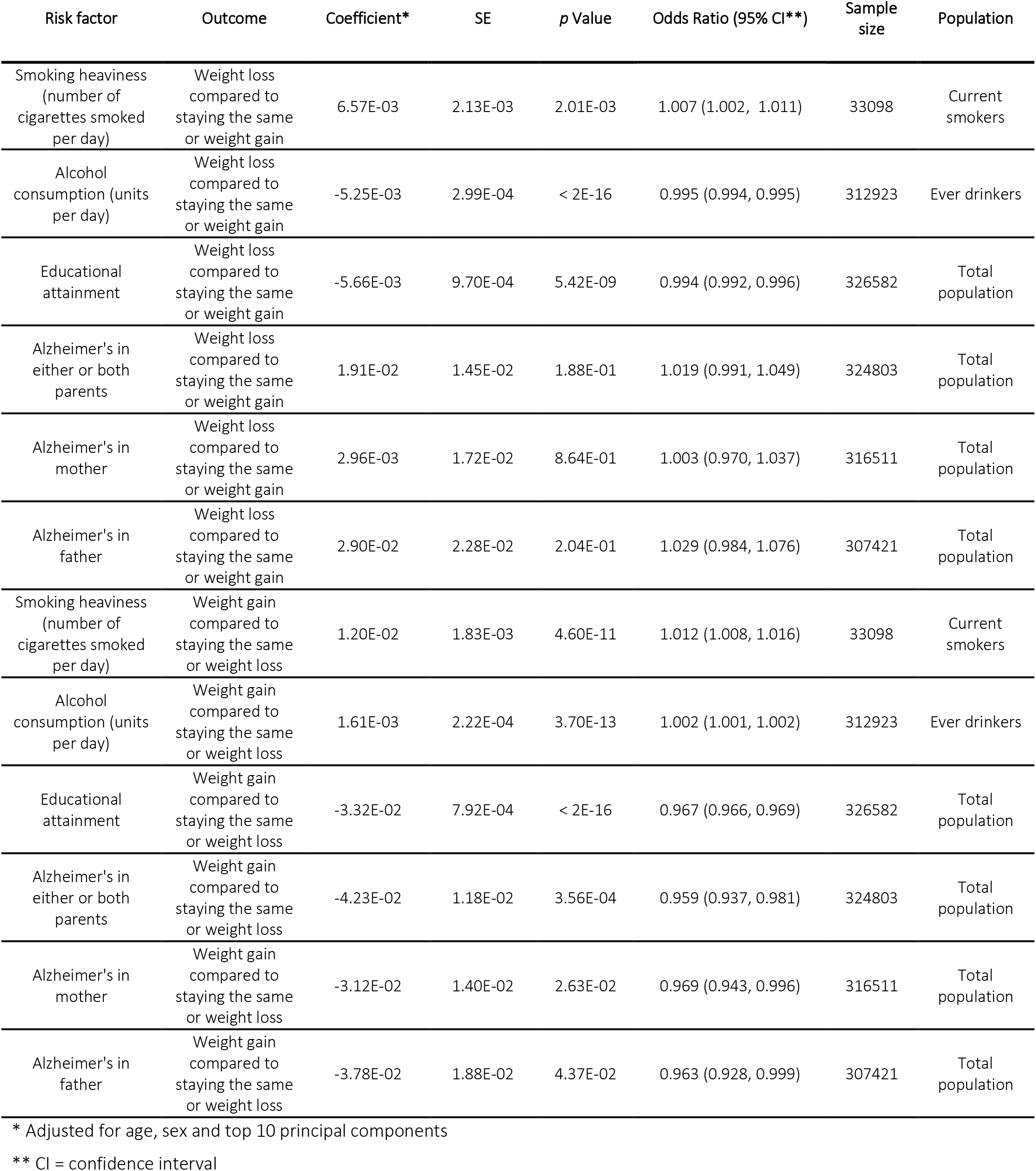
Conventional epidemiological estimates for risk factors on weight loss compared to staying the same weight and weight gain and weight gain compared to staying the same and weight loss during midlife.

**Table 2:** One-sample Mendelian Randomization estimates for risk factors on weight loss compared to staying the same weight and weight gain and weight gain compared to staying the same and weight loss.

| Risk factor | Outcome | Coefficient* | SE | p Value | Odds Ratio (95% CI**) | Sample size | Population |
| --- | --- | --- | --- | --- | --- | --- | --- |
| Smoking heaviness | Weight loss compared to staying the same or weight gain | 2.82E-02 | 0.011 | 8.12E-03 | 1.029 (1.007, 1.050) | 150799 | Ever smokers |
| Smoking heaviness | Weight loss compared to staying the same or weight gain | 2.81E-02 | 0.012 | 2.03E-02 | 1.029 (1.004, 1.053) | 116173 | Previous smokers |
| Smoking heaviness | Weight loss compared to staying the same or weight gain | 2.72E-02 | 0.022 | 2.21E-01 | 1.028 (0.984, 1.073) | 32284 | Current smokers |
| Alcohol consumption | Weight loss compared to staying the same or weight gain | -7.79E-04 | 0.005 | 8.78E-01 | 0.999 (0.989, 1.009) | 312923 | Ever drinkers |
| Educational attainment | Weight loss compared to staying the same or weight gain | -8.77E-03 | 0.005 | 7.04E-02 | 0.991 (0.864, 1.138) | 329531 | Total population |
| Alzheimer's genetic liability score | Weight loss compared to staying the same or weight gain | 2.26E-02 | 0.005 | 2.67E-06 | 1.023 (1.013, 1.033) | 329531 | Total population |
| Smoking heaviness | Weight gain compared to staying the same or weight loss | -3.91E-03 | 0.009 | 6.52E-01 | 0.996 (0.979, 1.013) | 150799 | Ever smokers |
| Smoking heaviness | Weight gain compared to staying the same or weight loss | 8.66E-03 | 0.010 | 3.76E-01 | 1.009 (0.991, 1.028) | 116173 | Previous smokers |
| Smoking heaviness | Weight gain compared to staying the same or weight loss | -5.03E-02 | 0.019 | 7.54E-03 | 0.951 (0.916, 0.987) | 32284 | Current smokers |
| Alcohol consumption | Weight gain compared to staying the same or weight loss | 1.08E-05 | 0.004 | 9.98E-01 | 1.000 (0.992, 1.008) | 312923 | Ever drinkers |
| Educational attainment | Weight gain compared to staying the same or weight loss | -4.77E-02 | 0.004 | < 2E-16 | 0.953 (0.946, 0.961) | 329531 | Total population |
| Alzheimer's genetic liability score | Weight gain compared to staying the same or weight loss | -2.76E-02 | 0.004 | 1.37E-11 | 0.973 (0.965, 0.981) | 329531 | Total population |

\* Adjusted for age, sex and top 10 principal components
\*\* CI = confidence interval

**Figure 2A:**
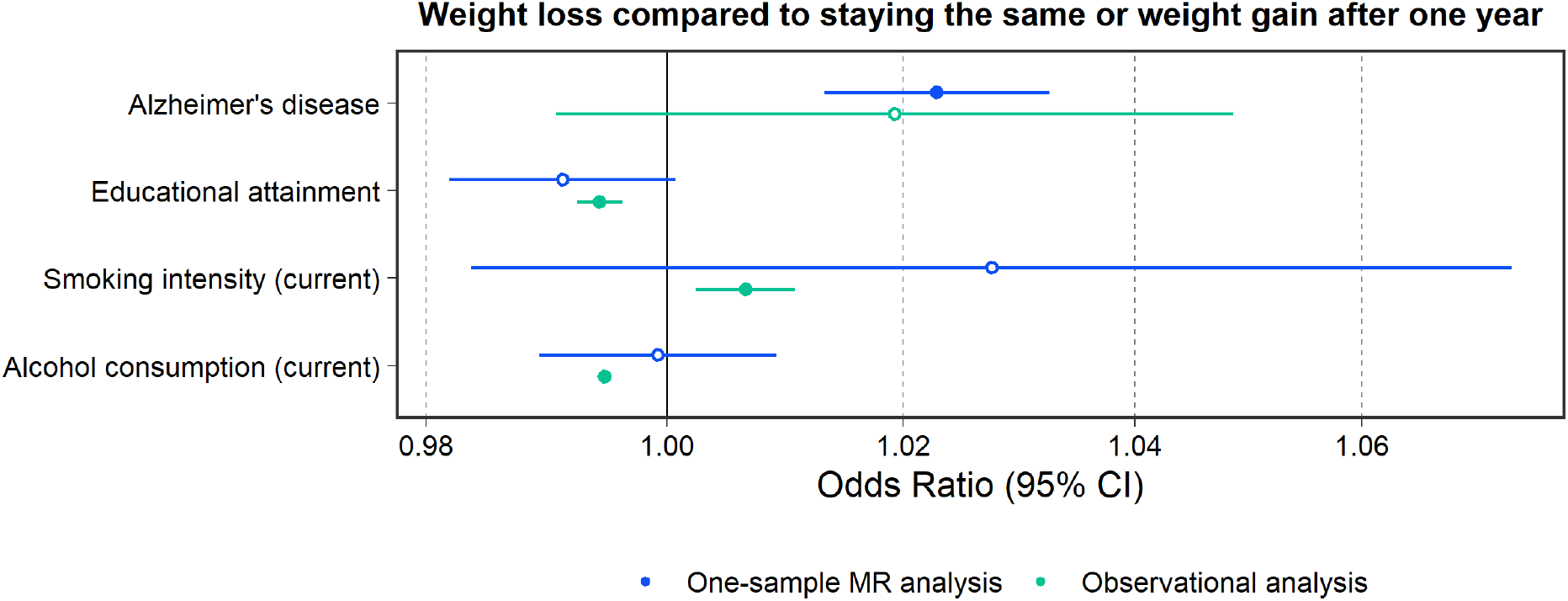
A forest plot with estimates for risk factors on weight loss comparing results from one-sample mendelian randomization (green) and associational (blue) analyses. The outcome variable, weight loss compared to staying the same and weight gain, comprises two tiers coded 0 (“No – weight about the same” and “Yes – gained weight”) and 1 (“Yes – lost weight”). Dots filled-in indicate some to very strong statistical evidence of an association and the exposure variable. Alzheimer’s disease is presented on the continuous liability scale in one-sample MR and coded 0 (“Alzheimer’s disease in neither parent”) and 1 (“Alzheimer’s disease in either or both parents”) in associational analyses.

When considering weight gain, conventional observational analyses indicated strong evidence that Alzheimer’s disease in either or both parents and educational attainment reduced odds of weight gain compared to staying the same or weight loss (OR, 95% CI: 0.967, 0.966 to 0.969, P<2.00×10^-16^ and 0.959, 0.937 to 0.981, P=3.56×10^-4^, respectively). There was also strong evidence that smoking intensity (number of cigarettes smoked per day) in current smokers (n=33 098) and alcohol consumption (units per day) in current drinkers (n=312 923) increased odds of weight gain (OR, 95% CI: 1.005, 1.002 to 1.008, P=0.002 and 1.002, 1.001 to 1.002, P=3.70×10^-13^, respectively).

In contrast to conventional observational estimates, MR analyses provided little evidence of an effect between alcohol consumption and weight gain in drinkers (OR, 95% CI: 1.000, 0.992 to 1.008, P=0.998) and the genetic instrument used for smoking heaviness suggested a decreasing effect of smoking heaviness on weight gain in current smokers (OR, 95% CI: 0.951, 0.916 to 0.987, P=0.008). In previous smokers, there was very little evidence that smoking heaviness had a genetically predicted effect on higher odds of weight gain (OR, 95% CI: 1.009, 0.991 to 1.028, P=0.376) (Table 1). The direction of the remaining effect estimates from one-sample MR were similar to conventional observational estimates for most exposures (Figure 2B).

**Figure 2B:**
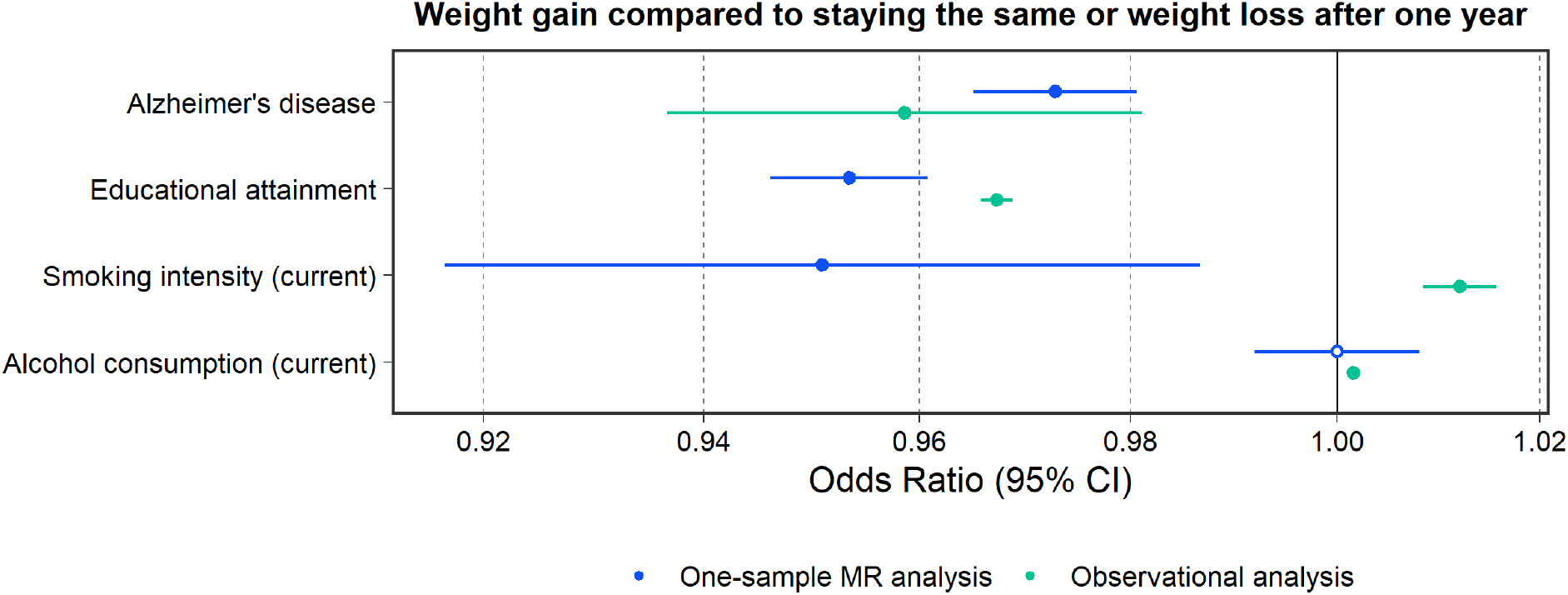
A forest plot with estimates for risk factors on weight gain comparing results from one-sample mendelian randomization (green) and associational (blue) analyses. The outcome variable, weight gain compared to staying the same and weight loss, comprises two tiers coded 0 (“No – weight about the same” and “Yes – lost weight”) and 1 (“Yes – gained weight”). Dots filled-in indicate some to very strong statistical evidence of an association and the exposure variable. Alzheimer’s disease is presented on the continuous liability scale in one-sample MR and coded 0 (“Alzheimer’s disease in neither parent”) and 1 (“Alzheimer’s disease in either or both parents”) in associational analyses.

### Collider bias

The directions of effects remain consistent with the main results, however, since including additional variables into IPW models reduced sample sizes, the precision of these estimates is likely to have been impacted which is reflected in corresponding confidence intervals (Table 3).

**Table 3:** Inverse probability weighted one-sample Mendelian Randomisation estimates for smoking intensity and alcohol consumption on weight change during midlife.

| Risk factor | Outcome | Coefficient | Standard error | p Value | Odds Ratio (95% CI**) | Sample size | Population |
| --- | --- | --- | --- | --- | --- | --- | --- |
| Smoking intensity | Weight loss compared to staying the same or weight gain | 0.030 | 0.007 | 1.88E-05 | 1.030 (1.017, 1.045) | 124989 | Ever smokers |
| Smoking intensity | Weight loss compared to staying the same or weight gain | 0.026 | 0.008 | 8.07E-04 | 1.026 (1.011, 1.042) | 106666 | Previous smokers |
| Smoking intensity | Weight loss compared to staying the same or weight gain | 0.068 | 0.030 | 2.47E-02 | 1.070 (1.009, 1.135) | 7669 | Current smokers |
| Alcohol consumption | Weight loss compared to staying the same or weight gain | -0.006 | 0.005 | 2.50E-01 | 0.994 (0.984, 1.004) | 201709 | Ever drinkers |
| Smoking intensity | Weight gain compared to staying the same or weight loss | 0.005 | 0.006 | 3.53E-01 | 1.005 (0.994, 1.071) | 124989 | Ever smokers |
| Smoking intensity | Weight gain compared to staying the same or weight loss | 0.012 | 0.006 | 6.55E-02 | 1.012 (0.999, 1.024) | 10666<br>6 | Previous smokers |
| Smoking intensity | Weight gain compared to staying the same or weight loss | -0.038 | 0.024 | 1.16E-01 | 0.963 (0.919, 1.009) | 7669 | Current smokers |
| Alcohol consumption | Weight gain compared to staying the same or weight loss | 0.001 | 0.004 | 8.08E-01 | 1.001 (0.993, 1.009) | 20170<br>9 | Ever drinkers |

\* Adjusted for age, sex and top 10 principal components
\*\* CI = confidence interval

### Validation

There was strong evidence that weight loss and weight gain during midlife conferred a heightened risk of all-cause mortality (OR, 95% CI: 1.393, 1.341 to 1.448, P<2.00×10^-16^ and OR, 95% CI: 1.201, 1.161 to 1.241, P<2.00×10^-16^, respectively) (Table 4). These results are consistent with previous studies (3–5).

**Table 4:**
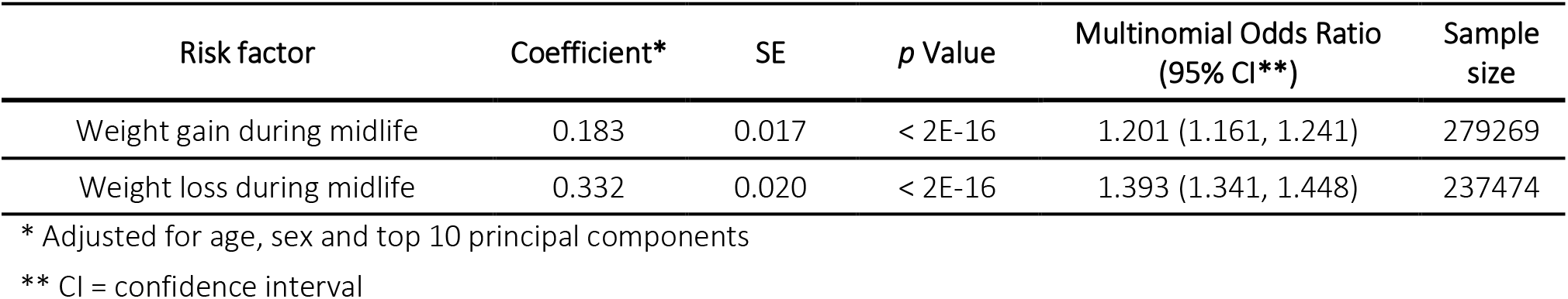
Observational estimates for weight gain and weight loss on all-cause mortality.

| Risk factor | Coefficient* | SE | p Value | Multinomial Odds Ratio (95% CI**) | Sample size |
| --- | --- | --- | --- | --- | --- |
| Weight gain during midlife | 0.183 | 0.017 | $< 2E-16$ | 1.201 (1.161, 1.241) | 279269 |
| Weight loss during midlife | 0.332 | 0.020 | $< 2E-16$ | 1.393 (1.341, 1.448) | 237474 |

\* Adjusted for age, sex and top 10 principal components
\*\* CI = confidence interval

### Sensitivity analyses

Findings from the two-sample MR weighted median and MR-Egger methods revealed similar trends to one-sample MR. This suggests little evidence that horizontal pleiotropy is driving our results, with the exception of the genetically predicted effect of alcohol consumption and educational attainment on weight loss, which did not support estimates from the original analysis. MR leave one out analysis showed that when ‘rs12972156’, located at the APOE locus on chromosome 19, is left out, the effect estimates for genetically predicted Alzheimer’s disease liability on weight change over a one-year interval in midlife remained very close to the original two-sample results (Supplementary Table S2). Results indicated similar trends within the group reporting no major dietary changes in the last 5 years, compared to the total population (Supplementary Table S3). Whilst not a direct indicator, we may infer, with caution, that weight change in this study is indicative of unintentional weight change, as opposed to intentional weight change.

## Discussion

In this study, conventional observational results indicated evidence of an association between educational attainment, Alzheimer’s disease family history, smoking intensity and alcohol consumption and weight change (gain and/or loss) over a one-year interval in midlife. The *directions* of effect within an MR setting were consistent with conventional observational estimates for educational attainment, Alzheimer’s disease liability, and alcohol consumption, though not smoking heaviness. Furthermore, the strength of these effects varied. IPW was used to account for non-random selection on smoking and alcohol status and indicated results were largely robust to collider bias.

Findings on educational attainment in our study were consistent with *a priori* expectations derived from previous studies. Specifically, evidence suggesting an association between educational attainment and lower odds of weight gain, which has been observed in studies conducted in high-income countries (16). Since educational attainment was also associated with reduced weight loss in our study, a known risk factor in clinical settings for various cancers and gastrointestinal diseases, for example, receiving higher education may consequently relate to reduced risk of mortality. In addition, results for educational attainment should partially be interpreted as a proxy for socioeconomic position and resource availability, as well as health and wellness promoting behaviours. Education influences weight-related actions such as diet and physical activity, thus affecting energy balance. If health behaviours of this kind persist, an imbalance has the potential to accumulate over time.

Alzheimer’s disease liability was a predictor of weight loss in our analyses. Importantly, using the APOE variant as a genetic instrument for Alzheimer’s disease liability may be less prone to misclassification than reported family history, which could explain the narrower confidence intervals when comparing conventional observational and MR estimates for this exposure. This result was supported using a two-sample MR leave one out analysis. Here we found removal of the ‘rs12972156’ genetic variant, located at the APOE locus, revealed the same direction of effect as our one-sample results. Whilst it is widely recognised that unintentional body weight loss is common in patients with dementia (17), the literature around whether weight loss may accelerate before the diagnosis of dementia is inconsistent and conflicting, further illustrating the presence of confounding in conventional epidemiolocal analyses (2). Our results are in line with previous research that shows that weight loss in midlife may come before a dementia diagnosis in later life (2, 39). Importantly, Alzheimer’s disease liability in this study in a conventional observational setting is based on family history data, whereas MR estimates are derived using genetic variants which may exert lifelong effects on disease risk. As such, it is challenging to directly compare the magnitudes of effect between these two types of analyses under the gene-environment equivalence assumption.

Conventional epidemiolocal analyses regarding alcohol consumption indicated that an increase in alcohol use was associated with weight *gain*. MR estimates, however, were much weaker. This may be in part due to our outcome measuring weight change over a one-year interval in midlife, assuming that on average, those that drink in this age group have been doing so for a considerable length of time. Thus, weight change as a result of drinking is likely to have had a substantial effect already. Conventional observational evidence of an impact within one-year is therefore potentially misleading, since this may be a result of confounding factors associated with alcohol consumption in midlife (40). Whilst conventional epidemiological analyses indicated an association between smoking intensity and weight gain over a one-year interval in midlife within current smokers, one-sample MR analyses were in line with the literature supporting an inverse relationship between current cigarette smoking and body weight (8, 41), suggesting evidence of an effect between smoking heaviness and reduced weight gain. In addition, having been a heavier smoker previously has been reported to lead to greater weight gain amongst previous smokers (41). Higher levels of BMI have also been reported to increase risk of smoking uptake as well as intensity of smoking (13). This highlights the need to combine weight control and smoking cessation strategies within interventions aimed at tackling these important public health concerns. To reiterate, careful consideration is required if stratifying a population into subgroups when conducting MR analyses, to evaluate when collider bias may have been introduced. We express the importance of adjusting for this using IPW (34).

Malignancies, non-malignant organic and psychiatric disorders constitute the main etiologic groups of unintentional weight loss (42). Weight loss is therefore often used clinically as an early indicator of the presence of disease. Our results, as well as findings from the Whitehall Study, a large cohort comprising male participants working in civil service offices aged 40-64 years (n=17,718), indicate that mortality is much greater in those that had reported weight loss over one-year compared to those that had not (4). Our results additionally show that weight gain is associated with mortality, though the effect is half the size of that of weight loss. Since both the UK Biobank and Whitehall Study consist of healthy populations due to the healthy volunteer and healthy worker effect, we expect these associations to be stronger in the general population.

### Strengths and limitations

Conventional epidemiological analyses have previously been conducted alongside MR to compare findings; however, this study is distinctive in that it assesses a time varying outcome; weight change over a one-year interval in midlife. This study also utilises additional sensitivity analyses through weighted median and MR-Egger methods in two-sample MR to strengthen the level of inference indicated in univariable results.

Important limitations exist in this investigation, however. Firstly, the outcome is a self-reported measure of perceived weight change over a one-year interval in midlife. Individuals with certain characteristics, including higher educational attainment, may be more likely to misreport their weight, indicating the potential for recall bias and thus differential misclassification (43). Secondly, the outcome used does not demonstrate weight change over a substantial period of time (i.e., more than one year) since the focus of this study is to examine recent and rapid weight change during midlife. Additionally, we do not have data on whether an individual changed their smoking or drinking throughout the year in which we examine their weight change. A further key limitation of this study is selection bias and thus limited generalisability. Participants in the UK Biobank are older, more likely to be female and less likely to live in socially deprived areas, compared to participants in nationally representative data sources (44). In addition, previous MR studies suggest that longer educational duration and older menarche relate to higher participation, whilst a genetic liability of neuroticism, Alzheimer’s disease and schizophrenia reduce participation (45). Finally, (i) selection on smoking and alcohol use, and (ii) stratification by smoking status, may have induced collider bias (34). Whilst IPW was conducted to account for this, including additional variables to generate weights resulted in smaller sample sizes and thus reduced power and precision in our IPW estimates.

## Conclusion

This investigation provides novel evidence of the effects of lifestyle risk factors, educational attainment, and Alzheimer’s disease liability on weight changes over a one-year interval in midlife. Results suggest that individuals who attended higher education may have more opportunity to help retain a steady weight during this stage in the lifecourse. At the same time, findings may have implications for early testing for Alzheimer’s disease, amongst other disease conditions, when weight loss is observed at midlife. The relationship between smoking intensity and weight change is complex, endorsing the notion that strategies to tackle weight control and smoking cessation should be combined.

## Data Availability

The data that support the findings of this study are all available from within the supplementary materials or publicly accessible from the resources cited.

## List of abbreviations

BMI: Body mass index
CI: Confidence interval
IPW: Inverse Probability Weighting
MR: Mendelian randomization
OR: Odds ratio

## Declarations

### Ethics approval and consent to participate

The UK Biobank study have obtained ethics approval from the Research Ethics Committee (REC; approval number: 11/NW/0382). The primary data from the UK Biobank resource that support the findings of this study are accessible upon application (https://www.ukbiobank.ac.uk/).

### Consent for publication

Not applicable.

### Availability of data and materials

The primary data from the UK Biobank resource that support the findings of this study are accessible upon application (https://www.ukbiobank.ac.uk/).

### Authors’ contributions

GMP conducted all analyses and drafted the manuscript. GDS and TGR conceived the study design, obtained funding, and supervised the project. JT, AG and JH provided support and statistical advice on analyses conducted. SF helped clean data and derived a summary variable. All authors helped refine the final version of the manuscript and approve with its submission.

## Acknowledgments

We would like to thank the UK Biobank study and participants who contributed to it (app #76538).

